# The Development of the Substance Use Disorder Screening Test (SUDST) according to the DSM-5

**DOI:** 10.1101/2022.05.30.22275768

**Authors:** Sukuma Saengduenchai, Sumnao Nilaban, Tanya Singtho, Apichart Ranuwattananon, Rasmon Kalayasiri

## Abstract

**Objectives:** A screening tool for substance use disorders is most needed for patient care and referral. The study aims to develop the Substance Use Disorder Screening Test (SUDST) to classify the severity of substance use disorders based on the Fifth Edition of the Diagnostic and Statistical Manual of Mental Disorders (DSM-5).

**Methods:** Eleven items of the SUDST, developed based on the DSM-5, were tested on 207 participants who were receiving treatment for methamphetamine use. Participants were interviewed with the SUDST, the screening test of the Ministry of Public Health Version 2 (V.2), the Mini International Neuropsychiatric Inventory (M.I.N.I) and were clinically diagnosed by attending psychiatrists.

**Results:** Cronbach’s alpha coefficient was 0.79. The SUDST was highly significantly associated with the M.I.N.I., V.2, and the clinical diagnosis (p < 0.001). Factor analysis showed three components: 1) preoccupation and loss of control 2) risky/harmful use and 3) impact to bio-psychosocial aspects. Of the total score of 11, the cut-off points to identify severe, moderate, and mild levels of risk were ≥7, 5-6, and 3-4, respectively with sensitivity = 72.7%-96.5% and specificity = 61.9%-88.7%.

**Conclusions:** The SUDST had high reliability and validity that could be used for screening risk for substance use disorders.

## Introduction

Substance use has caused problems in all continents around the world, affecting individuals, families, communities, and societies. In the South East Asian countries, the most common illicit substance use was cannabis, kratom, and methamphetamine (1-3), which the latter (commonly known as speed pill (yaba) and crystal meth (ice)) caused the most problem in the region. The Princess Mother National Institute for Drug Abuse Treatment (PMNIDAT), the first and largest treatment center for substance use disorders in Central Thailand and in the South East Asia with 600 in-patient bed capacities, reported statistics which showed nearly half of the patients having methamphetamine use disorder (4), which has been increasing each year from 40.26% in 2016, increasing to 46.39% in 2017, and to 54.35% in 2018. Most of the inpatients with methamphetamine use disorder in this treatment center had a history of psychiatric symptoms including anxiety or suspiciousness (91.2%), depression (86.8%), and hallucination (85.7%) (5).

An effective and user-friendly assessment tool for substance use disorders is most needed for patient care and referral to the allocated treatment settings for patient management. Treatment facilities for substance use disorder in Thailand has used The Ministry of Public Health’s Screening Test Version 2 (V.2), which was adapted from the World Health Organization (WHO)’s Alcohol, Smoking, and Substance Involvement Screening Test (ASSIST) (6, 7). The V.2 has been used to assess levels of risk of substance use for allocating persons who used substance to each level of the formal treatment settings in Thailand. Persons who had low risk from the screening test were usually referred to the outpatient treatment at the general, provincial, or district hospitals with service for persons who used substances while those with moderate risk were usually sent to the treatment program at the designated government locations in each province that was operated in collaboration between the Ministry of Public Health, Ministry of Defense, and Ministry of Justice, and Ministry of Interior (8). For people with high risk of methamphetamine use disorder as screened by the V.2 were usually sent to the inpatient treatment at the substance treatment facilities widely spread over four parts of Thailand.

In 2013, the American Psychiatric Association issued the Fifth Edition of the Diagnostic and Statistical Manual for Mental Disorder (DSM-5) (9) that view the substance use problems as a continuum of disorder from mild, moderate to severe substance use disorder. As such, the DSM-5 diagnosis for substance use disorders, especially methamphetamine, the most problematic illegal substance in the region, has never been tested or compared to the Thai V.2. In the study, we compared the DSM-5 criteria for substance use disorders with the current formal use of V.2, which is the main screening tool for allocating persons with illegal problems in Thailand to receive treatment by using the Substance Use Disorder Screening Test (SUDST), that had been newly developed based on the DSM-5 criteria symptoms. The SUDST was then tested in the study for validity and reliability with clinical diagnosis by clinicians and the Mini International Neuropsychiatric Interview (M.I.N.I.). The study aims to investigate the reliability and validity of this new screening tool for clinical substance use disorder based on DSM-5 criteria. The new instrument may be used accordingly with other personal and societal factors for allocating persons who use substance to get appropriate treatment for substance use based on their needs.

## Methods

Two-hundreds and seven male and female patients aged 13 years and over receiving outpatient service for methamphetamine use from the PMNIDAT and its satellite hospitals in the four regions of Thailand including Central (PMNIDAT or Thanyarak Pathumthani), North (Thanyarak Chiang Mai), Northeast (Thanyarak Khon Kaen), and South (Thanyarak Songkla) were interviewed between August 2017 to December 2018 by attending psychiatrists for DSM-5 substance use disorder diagnosis. All of them had at least 3 years experience in addiction treatment. The attending psychiatrists were blinded to the diagnosis made by the questionnaires used in the study. Risk and methamphetamine use disorder were obtained using the SUDST, the M.I.N.I. for methamphetamine use disorder and the V.2 by research nurses (with at least 5 years experience in addiction treatment). All interviewers attended a training class for M.I.N.I., SUDST, and V.2. Individuals with cognitive impairment or psychosis (e.g., history of being diagnosed with dementia, psychotic disorder) were excluded from the study. The study has been approved by the Ethics Committee of the PMNIDAT with study number: 59023.

The SUDST were developed from the DSM-5 criteria for substance use disorders (9). The test had 11 items for the respondent to answer yes or no to each question that correspond to what participants have experienced in the past 12 months of their substance use. The test was examined for content validity from 5 experts in the fields of addiction, behavioral science, and psychometric test. The Index of Item-Objective Congruence is 0.65. The screening tool was then adjusted per suggestion and tested in 30 subjects (10) with substance use. The language was then further adjusted based on suggestion from this initial field testing. The language-adjusted screening test was then tested for inter-rater reliability by two independent interviewers with the time interval not more than 1 week apart. The content of the screening test includes four aspects of addiction including impaired control of substance use, social impairment from substance use, risky use of substance, and the pharmacological aspect of addiction.

The V.2 is the screening tool for referral persons with substance use problems in Thailand. It was developed by the Ministry of Public Health (MOPH) and was known as “Screening Form – MOPH: Version 2” or V.2 in brief. The test was adapted from the World Health Organization (WHO)’s Alcohol, Smoking, and Substance Involvement Screening Test (ASSIST), comprising of 6 questions related to behaviors of substance use, the impact of substance use, and the concern from others about one’s substance use in the past 3 months. Each item has 5 answer options from never, only 1-2 times total, about 1-3 times per month, about 1-4 days per week, and 5-7 days per week (almost every day) The total score was categorized to three levels (e.g., mild, moderate, and severe) risk for substance dependence.

The M.I.N.I., Methamphetamine Section, Thai version was adapted for the diagnosis of DSM-5 Substance Use Disorder in the study by R.K. and colleagues. The original Thai M.I.N.I had two parts for substance diagnoses based on DSM-IV criteria to diagnose methamphetamine dependence (7 items) and methamphetamine abuse (4 items). In the current study, the item related to legal problem from substance use was excluded from algorithm of diagnosis and replaced by an item related to craving for substance in order to correspond to the DSM-5 criteria for substance use disorder. Having 2-3, 4-5, and 6 or more criteria mean having mild, moderate, or severe level of substance use disorders.

Data from all questionnaires were checked for completeness by a researcher. The total duration of data collection was 16 months. First we tested for reliability of the SUDST by using Cronbach’s alpha coefficient for internal consistency. Concurrent validity of the questionnaire for DSM-5 diagnosis for substance use disorder was tested by using contingency coefficient with the clinical diagnosis. In addition, construct validity was tested by using Factor analysis. Finally, the SUDST was tested to determine the cut off point for the risk of having substance use disorder as provided by clinicians’ judgment. Cut off points for differentiating levels of substance use disorders were analyzed by using area under the receiver operating characteristic (ROC) curve to have sensitivity and specificity values of the SUDST, the new screening tool for clinical substance use disorder based on DSM-5 criteria.

## Results

Table 1 shows demographics of the participants in the study. Of 207 participants, most were male (88.4%), single (60.9%), and graduated from high school (57.0%). The mean age was 29 years (SD = 8.3, Min = 14, Max = 53). Twenty-seven percent of the participants were unemployed and 67.1% had drug related offences in the past. Table 2 shows the substance use data. Most of the participants used methamphetamine and other illegal substances including cannabis and kratom (67.6%). About one-third (29.0%) used speed pills (yaba) only without any other illegal substance. The average daily amount of speed pills (yaba) consumed was 4 pills and the average duration of substance use was 7 years. Most individuals who used crystal meth (ice) did not know the daily amount in grams.

**Table 1.**
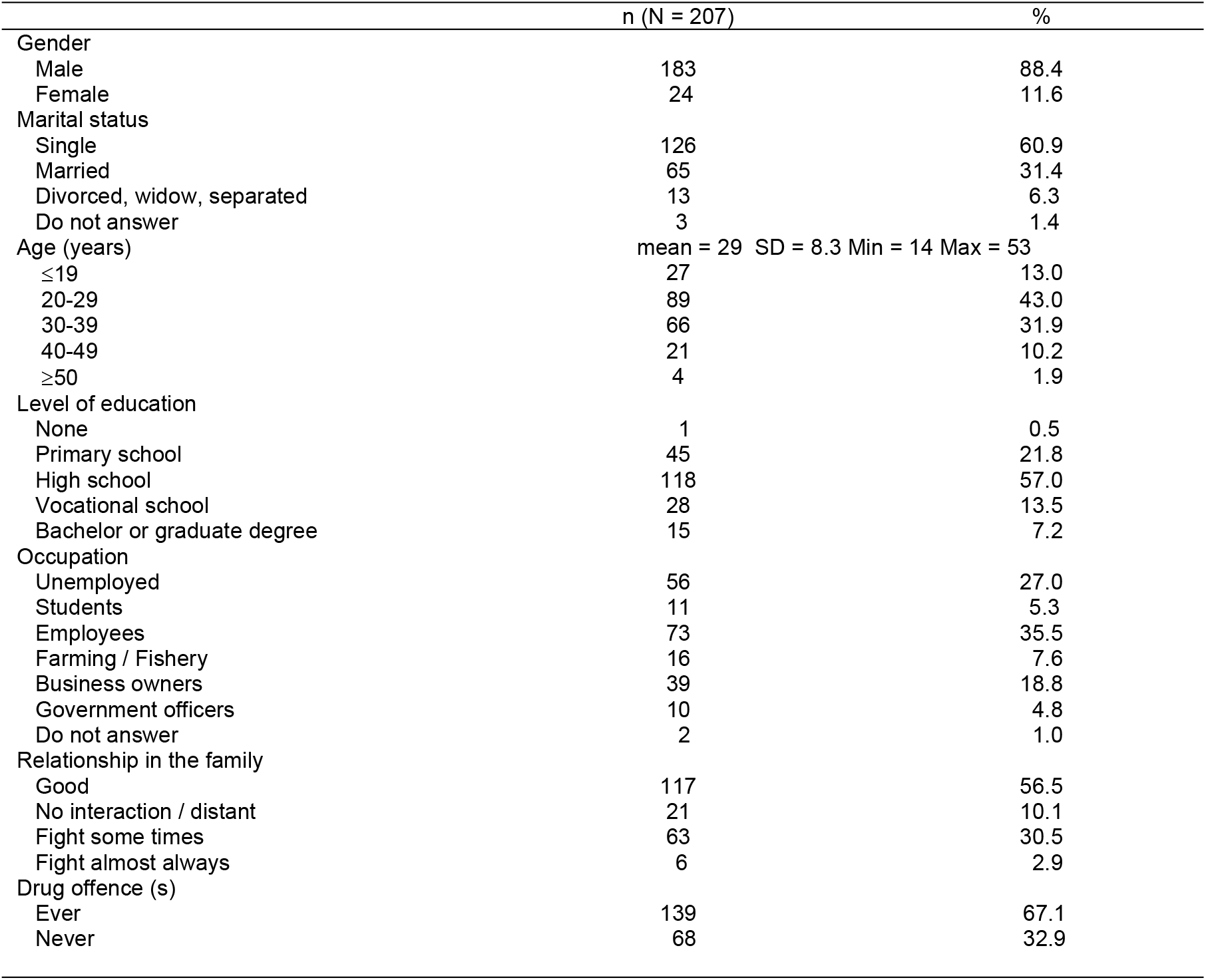
Demographic data of the participants in the study (N = 207).

**Table 2.** Substance and methamphetamine use data in the participants of the study.

|  | n (N = 207) | % |
| --- | --- | --- |
| Substance use |  |  |
| Speed pills (yaba) only | 60 | 29.0 |
| Crystal meth (ice) only | 3 | 1.5 |
| Speed pills and crystal meth only | 4 | 1.9 |
| Yaba/ice and other substances | 140 | 67.6 |
| Daily amount of yaba use (pills) | mean =4 SD = 5.1 Min =0.5 Max 48 |  |
| ≤ 5 | 143 | 72.9 |
| 6-10 | 20 | 10.1 |
| 11-15 | 4 | 2.0 |
| 15 and above | 2 | 1.0 |
| Did not answer | 29 | 14.0 |
| Duration of yaba use (years) | mean =84 SD = 74.4 Min =12 Max 576 |  |
| ≤ 5 | 104 | 52.5 |
| 6-10 | 45 | 22.7 |
| 11-15 | 25 | 12.6 |
| 16-20 | 15 | 7.6 |
| 20 or more | 4 | 2.0 |
| Did not answer | 5 | 2.6 |
| Daily amount of ice use (grams) |  |  |
| < 1 gram | 17 | 25.4 |
| 1 gram and more | 4 | 5.9 |
| Did not answer | 46 | 68.7 |
| Duration of ice use (years) | mean =36 SD = 49.9 Min =2 Max 240 |  |
| ≤ 1 | 10 | 14.9 |
| 2-5 | 10 | 14.9 |
| 6 or more | 2 | 2.9 |
| Did not answer | 45 | 67.3 |

Regarding reliability of the SUDST, Cronbach’s alpha coefficient is 0.79 which reflects good internal consistency of the instrument but Cohen’s kappa is only 0.45 for inter-rater reliability. Construct validity was tested by using Factor analysis of the 11 questions used in the questionnaire and shown in Table 3. Extraction method was used with the Principal component analysis by the Varimax rotation. Three factors were found including preoccupation / loss of control of use comprising of 4 questions, Risky or harmful use comprising of 3 questions, and the impact of use comprising of 4 questions. The correlation between each construct is in high level (more than 0.4). The factor loading could describe the variance at 51.8%. Specifically, Factor 1 could describe the variance at 32.3%, Factor 2 could describe at 10.1%, and Factor 3 could describe at 9.3%. Concurrent validity of the SUDST for Substance Use Disorder with other measures including clinical diagnosis by physicians and the M.I.N.I., and the V.2 were high by using contingency coefficient (p < 0.001) (Table 4).

**Table 3.**
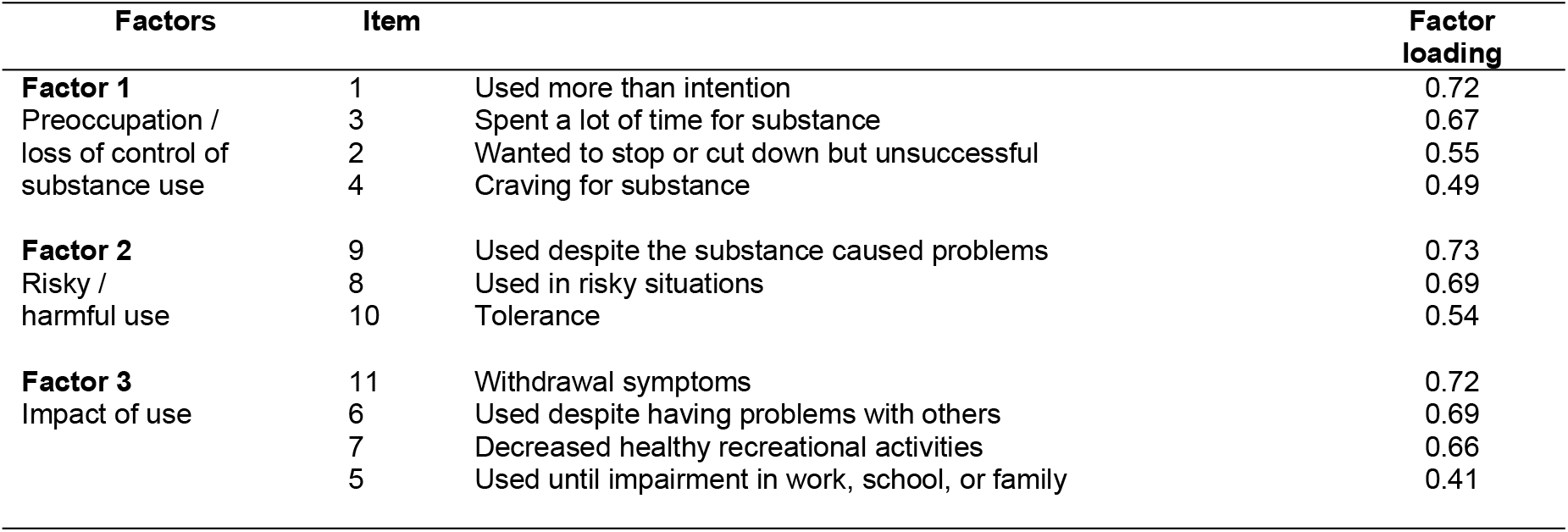
Factor loading of the 11 items of the Substance Use Disorder Screening Test (SUDST).

**Table 4.** Concurrent validity of the Substance Use Disorder Screening Test (SUDST) with the M.I.N.I., V.2, and clinical diagnosis.

| Measures | Coefficient | P-values |
| --- | --- | --- |
| Mini international neuropsychiatric Inventory (M.I.N.I) | .51 | <.001 |
| V.2 | .57 | <.001 |
| Clinical/physician diagnosis based on DSM-5 | .54 | <.001 |

Table 5 shows cut off points of the SUDST to determine level substance use disorder and categorize them into three groups based on clinical judgment including low, moderate, and high risk of substance use disorder according to the physician’s judgement. The score at 7 or higher had high sensitivity (77.9%) and specificity (72.7%) to determine the high risk group. The scores at 5 or 6 can define the moderate risk group (sensitivity = 88.7%; specificity = 61.9%) and the scores at 3 or 4 can define the low risk group (sensitivity = 96.5%, and specificity = 66.7%). The Area Under the Curve (AUC) is in good ranges (0.83 - 0.87).

**Table 5.** Cut – off point for the Substance Use Disorder Screening Test (SUDST) comparing to the clinical judgment based on DSM-5 criteria for substance use disorders.

| Level of substance use disorder | Cut-off point | Sensitivity | Specificity | Area under curve (95% CI) |
| --- | --- | --- | --- | --- |
| Severe | ≥7 | 77.9 | 72.7% | 0.83<br>(0.76-0.90) |
| Moderate | 5-6 | 88.7% | 61.9% | 0.87<br>(0.81-0.93) |
| Mild | 3-4 | 96.5% | 66.7% | 0.85<br>(0.69-1.0) |

## Discussion

The SUDST is a screening test based on DSM-5 symptom criteria for substance use disorders comprising of 11 questions regarding to the past 12 months experience of substance use with yes or no answers. The score ranging from 0 to 11. It is suitable for addiction researchers/specialists to evaluate people who use substance regarding to DSM-5 diagnosis of substance use disorder by using DSM-5 diagnostic criteria or to evaluate risk of substance use especially methamphetamine by using the new cut off point for clinical purposes such as allocation of persons to specific settings (i.e. outpatient, inpatient, long-termed rehabilitation) (8).

The reliability by internal consistency of the Thai SUDST instrument is in good level with Cronbach’s alpha coefficient = 0.79. Construct validity by Principal component analysis found 3 factors covered 51.8% of the variance including 32.3%, 10.1% and 9.3%, for Factor 1 (preoccupation / loss of control), Factor 2 (risky and harmful use), and Factor 3 (impact of use), respectively. Concurrent validity of the instrument is good when compared with three measures including M.I.N.I., V.2, and clinical diagnosis/judgment (P < 0.001). In addition to using the DSM-5 diagnostic criteria for substance use disorders, new scores of the instrument could make cut off points able to divide subjects into three categories comparable to the clinical judgment including high risk (score = 7 or more with sensitivity = 77.9% and specificity = 72.7%), moderate risk (score = 5 - 6 with sensitivity = 88.7% and specificity = 61.9%), and low risk (score = 3 - 4 with sensitivity = 96.5% and specificity = 66.7%) with the AUC in good levels (0.83 - 0.87).

In the study, all of the participants used methamphetamine in the past 12 months and 67% of them used it with other illegal substances. This proportion is consistent with the annual information that more than 50% of the patients with methamphetamine use disorders at the treatment centre used other illegal substances in the past 12 months (4). Using more than one substance increase harm and negative outcomes which might interfere with level of risk and result to different result of the screening (11). We did not exclude participants who used more than one substances in this study. However, including persons who used other substances than methamphetamine reflects the real world situation that would make the SUDST to be used generally.

Factor analysis of the SUDST revealed three components including preoccupation or loss of control, risky/harmful use, and impact of use. The result is consistent with other instruments related to measuring the severity or risk of substance. For example, Khon Kaen University-Volatile Use Disorder Identification Test (KKU-VOUDIT) (12), a ten-question questionnaire, comprised of three components including intoxication or impact of use, preoccupation or loss of control, and harmful use. Nevertheless, The DSM-5 diagnostic criteria for substance use disorder suggested four components of the disorder including impaired control, social impairment, risky use, and pharmacological component (9). In general, preoccupation or loss of control over substance use is usually considered to be the core symptoms of addiction by the definition of addiction that continuing to use or seek for substances despite knowing that it causes harm to oneself. Four items were in this core component to reflect the impaired control including 1) using substance more than intended 2) unable to stop or cut down substance use 3) spending a lot of time using/interacting with the substance and 4) craving for substance.

The pharmacological aspect has not appeared as a component in the study. Specifically, the two questions reflecting the pharmacological aspect of substance use disorder (e.g., tolerance and withdrawal) were each either included into the component of risky / harmful use or in component of the impact of use. Regarding the items for the component of risky/harmful use, they include 1) using substance in a risky situation (i.e., driving, operating machinery) 2) using substance despite knowing it causes physical, psychological, or other problems and 3) increasing the amount of substance to get the same effect or experiencing reduced effect while using the same amount of substance. It is understandable that tolerance of a substance especially the question asking about increasing the amount of substance in order to get the same effect may be viewed as risky/harmful use by the persons who use substance.

Likewise, having withdrawal symptoms from methamphetamine, including fatigue, hypersomnia, irritable or depressed mood may be viewed as negative impacts of substance use and are included in the component of impacts of use. The other items in this component include 1) regular use of substance until having impaired function for work, study, or family 2) using substance despite it caused interpersonal relationship (i.e., domestic violence, physical or verbal fights) and 3) reduced social or recreational activities due to substance use.

Several limitations of the study deserve to be mentioned. The study took place in addiction treatment settings so the most of the participants mostly had problems from substance use and might not reflect those with milder level of substance use. Further study should test the instrument in primary health care where more persons with mild level or use without any level of substance use disorder would be included. In addition, larger sample size would give more power to test the instrument. People who use other substances including illegal substances (i.e., cannabis, kratom, opioids) or legal substances (i.e., alcohol, tobacco) should be recruited to test the reliability and validity of the instrument.

By using the SUDST, as well as the risk assessment by the V.2 which has already been formally used, the severity of the disorder is divided into three levels of risk including mild, moderate, and severe. By doing so, substance use disorder is viewed as a spectrum and not a binary of disease (e.g., abuse or dependence). This work is in agreement with the ASSIST (7) and the DSM-5 diagnostic criteria (9) (which is the basis of the SUDST). The SUDST cut-off point for the mild level of the risk for substance use disorder has 97% sensitivity, making the instrument a good candidate for screening persons with risky behavior of substance use to the various level of severity and referral to an appropriate setting at the beginning stages of the diseases. Therefore, the SUDST is an option to be used in health care setting to differentiate the severity of substance use.

## Data Availability

Data will be held in a public repository, https://cads.in.th/cads/.

## Acknowledgements

We would like to thank Viroj Verachai, M.D. for facilitating the process of the study. We would also like to thank all participants for their time and efforts. R.K.’s research career is supported by the Centre for Addiction Studies (supported by The Thai Health Promotion Foundation) and the R01 DA037974 and the D43 TW009087/TW/FIC NIH HHS, US.

